# Current forecast of COVID-19: a Bayesian and Machine Learning approaches

**DOI:** 10.1101/2020.12.11.20231829

**Authors:** Kernel Prieto

**Affiliations:** Instituto de Matemáticas, Universidad Autónoma de México, México

**Keywords:** Bayesian inference, machine learning, COVID-19, contact tracing, data-driven

## Abstract

We address the estimation of the effective reproductive number *R*_*t*_ based on serological data using Bayesian inference. We also explore the Bayesian learning paradigm to estimate *R*_*t*_. We calculate *R*_*t*_ for the top five most affected principal regions of Mexico. We present a forecast of the spread of coronavirus in Mexico based on a contact tracing model using Bayesian inference inspired in a data-driven approach. We investigate the health profile of individuals diagnosed with coronavirus in order to predict their type of patient care (inpatient or outpatient) and survival. Specifically, we analyze the comorbidity associated with coronavirus using Machine Learning. We implemented two classifiers, the first one, to predict the type of care procedure a diagnosed person with coronavirus presenting chronic diseases will obtain: outpatient or hospitalized. Second one, a classifier for the survival of the patient: survived or deceased. We present two techniques to deal with these kinds of unbalanced dataset related with outpatient/hospitalized and survived/deceased cases, occurring in general for these type coronavirus datasets in the world, in order obtain to a better performance for the classification.

## 1 Introduction

Different mathematical models for disease transmission have been proposed to predict and control disease spread since the emerging and re-emerging of infectious diseases represent a major threat to public health and may cause big economic and social losses. Vaccination is the principal control measure for reducing the spread of many infectious diseases [32, 37]. Some recent epidemics of H1N1, Ebola, MERS-CoV needed strong government interventions for fast eradication [12]. Based on past pandemics, scientists had worn that another pandemic could strike at any moment. Therefore, a big effort to study the impact of control measures to eradicate the outbreak of an epidemic has been done for taking an immediate response for a possible influenza pandemic crisis [38]. Mathematical models include compartmental epidemic models, which are deterministic systems of ordinary and partial differential equations or stochastic difference equations [14]. For some diseases such as influenza, typhoid fever, anthrax, diphtheria, tetanus, cholera, hepatitis B, pertussis, pneumonia, and coronavirus the process of transmission between individuals take place because an initial inoculation of a small amount of pathogen units. Then, the pathogen reproduces fast within the host during a period of time, called incubation time. During this period, pathogen affluence is enough low for activating the transmission to other susceptible [36]. Many mathematical models assume that the disease incubation is negligible, once a individual is infected, i.e., this infected individual becomes infectious instantaneously. A compartmental model based on these assumptions are named SIR or SIRS [47], depending on whether the acquired immunity is permanent or temporal. For viral infections such as rubella, and measles, the infected individual acquires permanent immunity. However, many diseases, such as influenza, typhoid fever, anthrax, diphtheria, tetanus, cholera, hepatitis B, pertussis, pneumonia and coronavirus have an incubation (latent) period of time before the hosts become infectious [7]. Also, diseases with long immune period include polio, chicken-pox, whooping cough, smallpox and dengue fever. In order to take into account this incubation period of the disease, another population compartment, named exposed class, *E*, is incorporated into these type of models, SIR and SIRS. Then, a susceptible individual who has been just infected, first goes through the exposed class during a incubation period of the disease, after that, the exposed individual becomes infectious. The resulting models are of SEIR or SEIRS type. We point out that there exists more literature on SIR and SEIR models than SIRS and SEIRS models, i.e., those which permanent immunity is not assumed. We refer the reader to [30, 46, 63] for references on SEIRS models and [6, 36, 39, 40, 42, 43] for references on SEIR models.

Numerous efforts to forecast and mathematical control models for disease transmission have been proposed since the re-emerging of the coronavirus named SARS-CoV-2 [4, 21, 26, 28, 44, 61, 64]. The first coronavirus outbreak, named SARS-CoV, where SARS stands for Severe acute respiratory syndrome, caused an pandemic with different incidences in the 29 countries around the world. A Bayesian compartment (SEIR: Susceptible, Exposed, Infected and Removed) model was presented to study the spread of the first coronavirus in 2002 [50]. The mean incubation period was 5.3 days (%95 Credible Interval 4.2 − 6.8 days), which is close to the latter coronavirus mean incubation period, reported as 5.1. Also the reported mean recovery period, from symptom onset to recovery, was 21 days (%95 Credible Interval 16 − 26 days), which is higher compared to the second coronavirus recovery period, reported around 14 days. A social distance as a control strategy of SARS was explored in [23]. The basic and effective reproductive numbers of SARS-Cov were estimated in [45]. Also, a spatiotemporal analysis of SARS-CoV was presented in [16]. We point out that other type of coronavirus emerged in 2015 in the Republic of Korea, named Middle East Respiratory Syndrome Coronavirus (MERS-CoV). After 17 years of the first apparition of SARS-CoV (November, 2002), another virus strain has emerged, called SARS-CoV-2. Since the second coronavirus outbreak, also named COVID-19, in Wuhan City in December of 2019, many attempts to predict the dynamics of coronavirus pandemic have been presented, some with a Bayesian inference approach [10, 20, 26]. A wide range of predictions have been presented in model calibrations using confirmed-case data since the nonidentifiability in theirs models [56].

The remainder of this paper is organized as follows. Section 2 describes some formulations to estimate the basic and the effective reproductive numbers, *R*_0_ and *R*_*t*_. Section 3 describes the mathematical formulation of the contact tracing model for coronavirus disease and the Bayesian inference framework to predict the dynamics its spread. Section 4 describes a clinical analysis of data set [1] using Machine Learning. Each section presents the mathematical framework and numerical results. Discussion and conclusions are presented in the last section 5.

## 2 Estimation of reproductive numbers *R*_0_ and *R*_*t*_

The basic reproductive number of a disease, *R*_0_, is the expected number of secondary infectious cases generated by an average infected person in an entirely susceptible population. The reproductive number *R*_0_ quantifies how many susceptible persons are on average infected by one infected person. If one infected person infect on average more than other person, then there exists an epidemic outbreak, in other words, the diagnostic infected people grow exponentially. Thus, *R*_0_ = 1 is the threshold value of the spread or decline of an epidemic. Estimation of *R*_0_ can be done in two ways, either by inferring it from observed case numbers, or by following infection chains step by step. Here, we will discuss the former. Estimating *R*_0_ using diagnosed cases can be done by some approaches as follows. Taking into account a latent period of the disease, i.e. during this stage, pathogen abundance is too low for active transmission to other susceptible hosts, although the pathogen is still present. The SEIR model takes into account this latent period. Thus, the COVID may be modeled with a SEIR model. The disease-free equilibrium point for *I* is given by

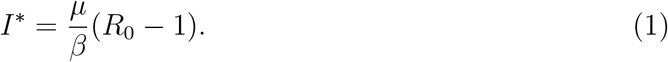

Doing an equilibrium analysis similar to the Box 2.4 of [36], the eigenvalues at the disease-free equilibrium allows us to describe the increase in prevalence during the invasion phase:

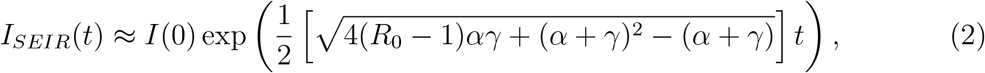

where *I* is the number of infected people, *α* is the rate the latent individuals become infectious and *γ* is the rate that infectious individuals become recovered and the reproductive number *R*_0_ is

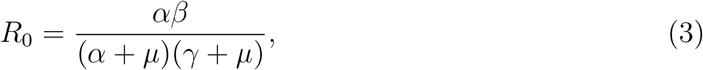

which is the product of the transmission coefficient(contact rate) *β*, the probability for the infected fraction becoming infective, *α/*(*α*+*µ*), and the average infectious period 1*/*(*γ*+*µ*), here *µ* is the birth rate. We have ignored the natural death rate to obtain (2) since we will use this approximation considering only the early stage of the epidemic and to simplify the equation. Next, we estimate *R*_0_ within the first 65 days of the epidemic using the least squares method. Here, we used the function *least* _*squares* from scipy Python package with argument *loss=soft*_*l1*. Using the parameter values *α* = 1*/*5.1, *α* = 1*/*14, *µ* = 1*/*(75×365), we obtain the estimation of *R*_0_ equal to 2.6232. Figure 1 show the fit of the early pandemic (65 days).

**Figure 1:**
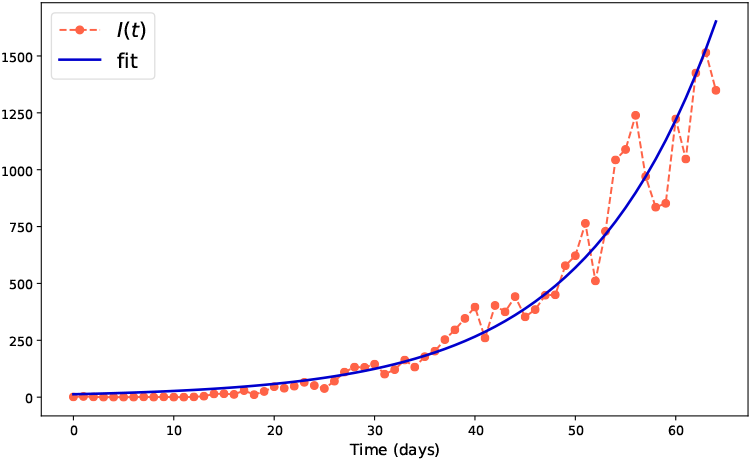
Fit of the early outbreak using (2)

Next, we will use the mean age at infection. An indicator of prevalence of an epidemic is the host’s mean age at infection, *A* (if the infected individual does not obtain immunity, then one would use the mean age at first infection). Calculating the mean time an individual remains susceptible, i.e., the mean time from birth to infection, the average period spent in the susceptible class is approximated by the inverse of the force of infection, 1*/βI*^*∗*^. Upon substituting for *I*^*∗*^ form equation (1), we obtain an expression for the mean age at infection:

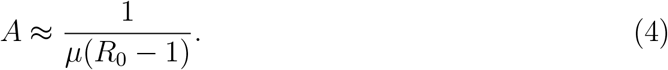

The above equation can be rephrased as *R*_0_ − 1 = *L/A*, where *L* is the host’s life expectancy. Using the host’s life expectancy of Mexico [2] *L* = 75.41 (both sexes), and calculating the mean age at infection *L* = 45.17 from data set [1], we estimate *R*_0_ using (4) as *R*_0_ = 2.66. Another alternative for calculating *R*_0_ is to use the package [53] written in R language. Authors in [3] have reported the value of *R*_0_ for the outbreak in the whole Mexico country as *R*_0_ = 2.7 (2.5, 3.2) and 2.3 (2.0, 2.6) for the outbreak the exponential growth and maximum likelihood method, respectively, using this package.

In contrast to *R*_0_, the effective reproductive number, *R*_*t*_ measures the number of secondary cases generated by an infected person once an epidemic is underway. We present an approach to estimate the effective reproductive number *R*_*t*_. This approach uses the seroprevalence data via the equilibrium point relation for susceptibles *S*^*∗*^ = 1*/R*_0_, i.e., *R*_0_ is estimated as the inverse of the inverse of the proportion of our sample that are seronegative (susceptible). This sample may not represent the entire population, because the level of seroprevalence is expected to increase with age. Therefore, one may use the age-dependent nature of the likelihood of being susceptible. For an individual of age *a*, the standard *SEIR* model predicts that the probability an individual is still susceptible is given by *P* (*a*) ≈ exp(−*aµ*(*R*_0_ − 1)). Thus, knowing the ages of the serological sample, we can construct the likelihood that the data comes from a disease with a particular *R*_0_ value, and then find the *R*_0_ that maximizes this likelihood. If we have *n* individuals who are susceptible (seronegative) of ages *a*_1_, …, *a*_*n*_, and *m* individuals who are seropositive at ages *b*_1_, …, *b*_*m*_, then the likelihood is:

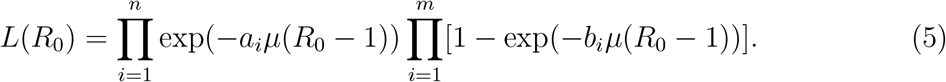

We have used the package *t-walk* [24] to estimate *R*_0_ maximizing the likelihood *L* of (5). The serological information [1] could be thoght to be biased since the tests done in Mexico have not been randomly, they have been programmed once a individual presents symptoms of coronavirus, but the results indicate that the sample scan has covered a huge range of age. The average age of seropositive is equal to 45 years and its standard deviation around 16 years. As an alternative to the approach described above, we applied the Bayesian Learning paradigm (BLP) [29] for the the time series of serological data set *y*. Assume we observe the first set of serological data at day 1, noted *y*_1_, and calculate the posterior density, *π*_*P*_ (*θ*|*y*_1_), from the likelihood function (5) along with the specific prior:

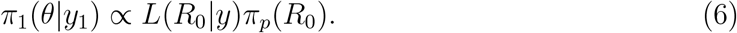

Next, we observe a second data at day 2, noted *y*_2_, independent of the first set but from the same data-generating process. To estimate the posterior *π*_2_(*θ*| *y*_2_, *y*_1_), we consider the previous posterior as a prior and proceed to calculate using a likelihood density from the second data, thus, instead of using all the data as it was arrived at once at a specific day *t*, we update the posterior *L*(*R*_0_|*y*_*t*+1_, *yt*) considering the posterior of one day before, noted *π*(*R*_0_ | *y*_*t*_), as a prior of the current day and repeat it until we arrive at the last day of our estimation:

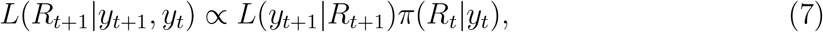

in other words: “Today’s posterior is tomorrow’s prior” [58]. Although, using the BLP is an approximation of the above approach, the BLP represents an enormous advantage considering a large scanned population during a pandemic, e.g., Mexico currently has tested more than one million of people. Without following the BLP, the computational time consumption is around two days (without splitting dates and saving the results). We point out that *t-walk* is not parallelized at the moment. The computational time consumption using the BLP is around 90 mins for the country and less than an hour for the top five most affected states in Mexico. We start formula (5) from March 21st in order to have enough sample size, and finishes on August 16th. Figure 2 shows the histogram of the seropostive and seronegative ages of tested people in Mexico. We can see in this figure that this histogram is similar to a Normal distribution histogram. Figure 3 shows the Effective reproductive number for the top five most affected regions, it starts on March, 21st and finishes on August 16th. The transparent bands represent the spread of the posterior distribution of (5). Figure 4 shows the Effective reproductive number for the top five most affected regions, it starts on March, 21st and finishes on August 16th using the Bayesian Learning Paradigm, it starts on March 21st and finishes on August 16th. The transparent bands represent the spread of the posterior distribution of (5). Figure 5 shows the Effective reproductive number for the whole Mexico country, it starts on March, 21st and finishes on August 16th. The transparent bands represent the spread of the posterior distribution of (5). Another alternative to estimate *R*_0_ is recently presented in [59], in this work. it is considered also the imported cases.

**Figure 2:**
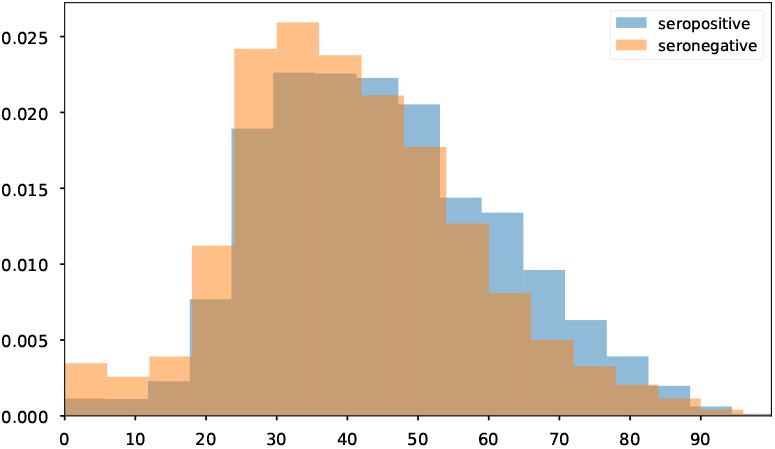
Histogram of the seropostive and seronegative ages of tested people

**Figure 3:**
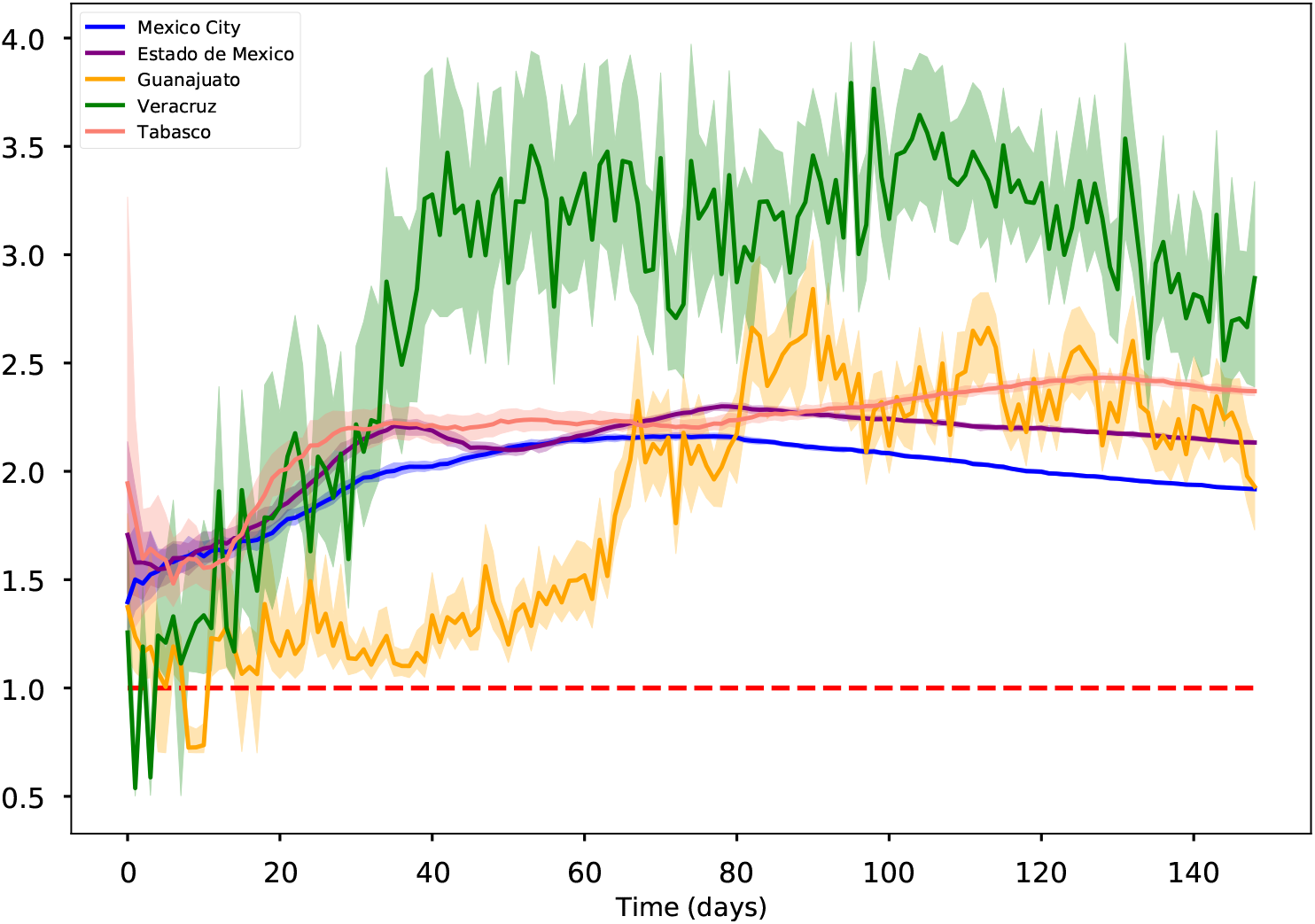
Effective reproductive number for the top five most affected regions, it starts on March, 21st and finishes on August 16th. Blue solid line is the median estimate and the transparent bands represent the 95% Highest-Posterior Density (HPD) interval

**Figure 4:**
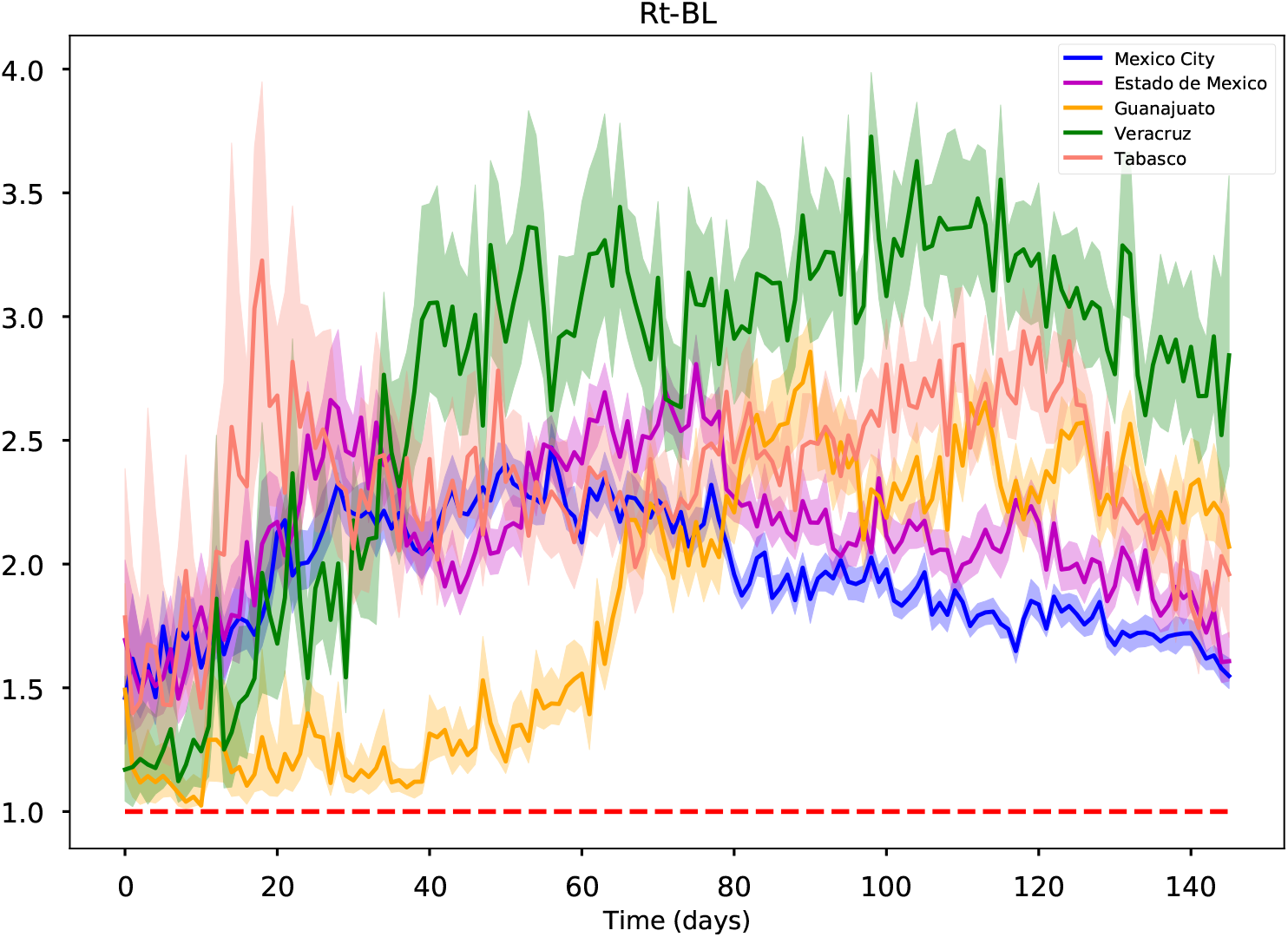
Effective reproductive number for the top five most affected regions using the Bayesian Learning Paradigm, it starts on March, 21st and finishes on August 16th. Blue solid line is the median estimate and the transparent bands represent the 95% Highest-Posterior Density (HPD) interval

**Figure 5:**
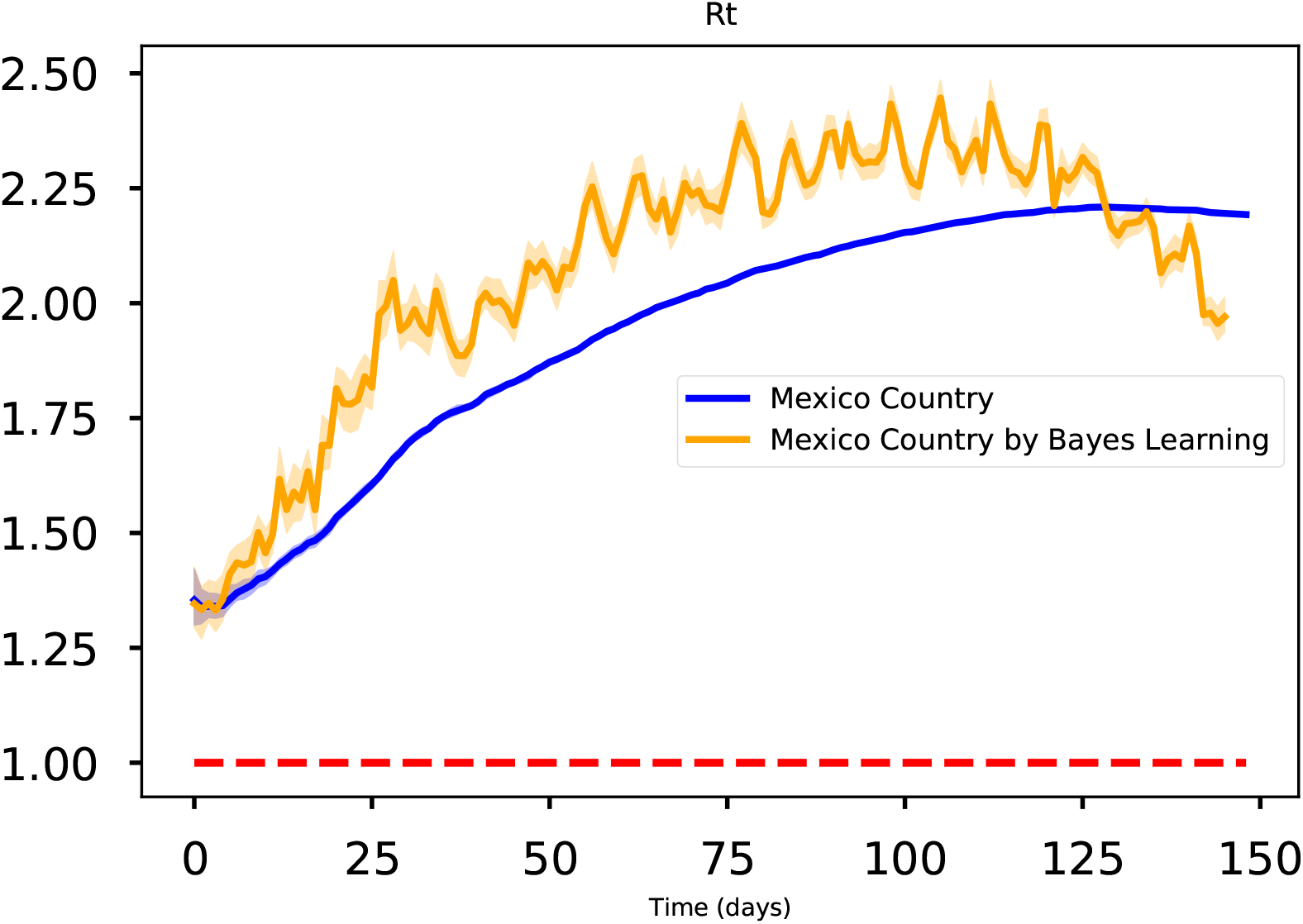
Effective reproductive number for the whole country, it starts on March, 21st and finishes on August 16th. Blue solid line is the median estimate and the transparent bands represent the 95% Highest-Posterior Density (HPD) interval

## 3 Bayesian forecasting

### 3.1 Model formulation

Effective Vaccination [17], early detection, proper treatment, isolation, quarantine, educational campaign are some control strategies to wane infectious diseases. With the aim to study the effect of contact tracing in the propagation of an infectious disease, we formulate a contact tracing model. Here, it is assumed that the disease transmits horizontally, i.e., vertical transmission is neglected. The horizontal transmission can occur either by direct contact, e.g, touching, licking, biting, or by indirect contact with no physical contact, e.g, vectors or fomites.

The frameworks ‘Susceptible-Infectious-Removed’ (SIR) and ‘Susceptible-Exposed-Infectious-Removed’ (SEIR) have been used in most current studies of COVID-19 transmission dynamics. Inspired in a full data-driven approach, we have tried to use all the reliable data available for forecasting the spread of the coronavirus disease, keeping in mind that simple model may fit better than complex ones [56]. Next, we formulate a mathematical model considering isolation due to contact tracing as suggested in [36] and the models proposed in [3, 23, 56]. This model analyzes the significance of isolate the probable infected individuals. The total population *N*, is divided in the following seven epidemiological classes SsEIQR: susceptible *S*, suspects (susceptible quarantined) *s*: people who have had contact with an infectious person or with someone who had contact with an infectious person), exposeds *E*, people who have contracted the virus but are not yet infectious, the undetected infectives *A*, asymptomatic people, sick people reported in quarantine *I* (these individuals are isolated at home or in the hospital), recovered people *R*, and the last state variable *P* denoting the deceases by coronavirus. We assume the disease transmission rate, *λ*, is decomposed in two parts, the disease transmission rate by symptomatic people and by asymptomatic people; *λ* = *β*_*a*_ + *β*_*s*_. We assume that a fraction *q* of the contacts whom infected individuals have had recently, are sought and isolated. We model contact tracing by forcing a fraction *q* of those who have recently had contact with an infectious individual to be quarantined where they will spend an average 1*/τ* days. importantly, we assume that these individuals are quarantined before they have a chance to generate any subsequent infection. Because of this latter assumption, contact tracing does not need to be recursive. The parameter *α*^−1^ and *γ*^−1^ represents the mean latent period and the recovery period, respectively. The parameter *ρ* represents the proportion between the symptomatic class and the asymptomatic class. Finally, the parameter *σ* denotes the death rate by the disease. Our model reads as follows

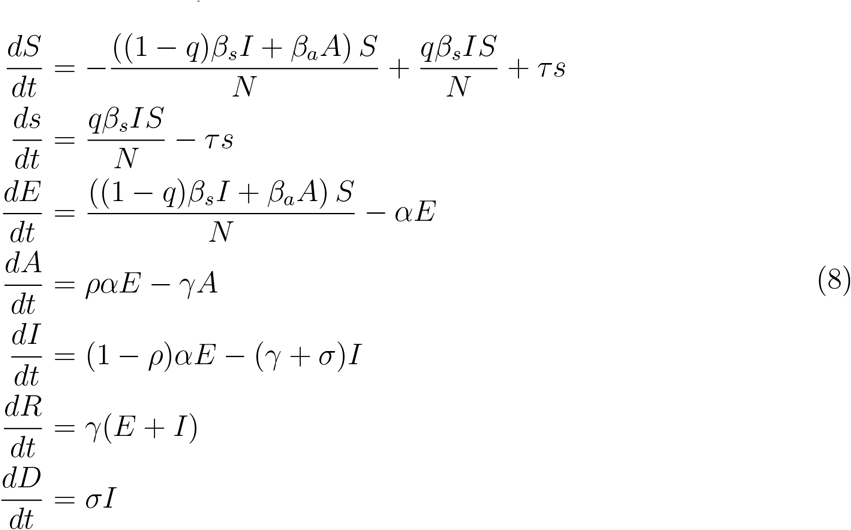

The total population *N* (*t*) is determined by *N* (*t*) = *S*(*t*) + *s*(*t*) + *E*(*t*) + *A*(*t*) + *I*(*t*) + *R*(*t*) + *D*(*t*). We point out that a more complex model is suggested in [36], considering stages in the exposed and infectious compartments but considering to decompose the force of transmission *λ*. A contact tracing model to explore for future work is in [27] where propose a very interesting and robust force of transmission *λ* dependent of time and with a delay. A sensitivity analysis shows that *λ* is the highest sensible parameter in this kind of compartment models, therefore, it is very important to select this parameter adequately. Another interesting options of contact tracing models are in [15]. A robust review of contact tracing models is in [41] and quarantine models in [34]. A detailed mathematical analysis of type of SEIR models are in [13, 33].

### 3.2 Parameter estimation

For the parameter estimation we used the daily updated data [1]. From the mathematical point of view, the parameter estimation of a system of ordinary differential equations is regarded as an inverse problem. Fitting curve or estimation the parameters of a model is considered an inverse problem from the mathematical point of view. Typically, an optimization method, e.g., the Landweber in [5, 18, 54, 55, 60], or faster methods such as the Levenberg-Marquardt or Conjugate Gradient methods, and regularization techniques, such as Tikhonov, Sparsity or Total Variation, are used to solve this inverse problem. In this manuscript, we used Bayesian inference to solve the inverse problem since it is a tool which combines uncertainty propagation of measured data with available prior information of model parameters. Also, it is numerically more stable approach than classical methods, since classical methods rely on the starting parameter point must be relatively close to the true one, otherwise the solution obtained corresponds to a local minimum. Moreover, classical methods gives only a point estimate solution instead of a band of the solutions using Bayesian inference, i.e., in a Bayesian framework, one works with credible intervals. Some references of using Bayesian inference are in [3, 8, 9, 10, 11, 14, 17, 21, 22, 31, 48, 62]. A Bayesian framework to model the spread of the first coronavirus,i.e., SARS-CoV, was presented in [50]. Using Bayesian inference, solutions of the inverse is obtained from the posterior distribution of the parameters of interest, an a solution of interest is obtained using the Maximum a Posterior, called MAP. This MAP gives the parameter value for which the posterior density is maximal. Also, one can calculate the median and quantiles from this posterior sample. As already mentioned, the Bayesian framework provides a natural and formal way to quantify the uncertainty of the quantities of interest. Denoting the state variable *x* = (*S*(*t*), *s*(*t*), *E*(*t*), *I*(*t*), *Q*(*t*), *R*(*t*), *P* (*t*)) ∈ (*L*^2^([0, *T*])^*n*^, i.e., *n* denotes the number of state variables, here *n* = 7, and the parameters *θ* = (*β, q, δ, α, γ, σ, s*(0), *E*(0), *I*(0), *Q*(0)) ∈ ℝ^*m*^, i.e., *m* denotes the dimension number of parameters to estimate, here *m* = 10, we can write the model (8) as the following Cauchy problem

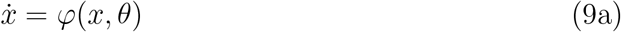

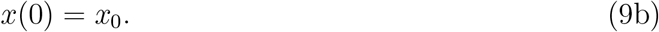

Problem (9), defines a mapping Φ(*θ*) = *x* from parameters *θ* to state variables *x*, where 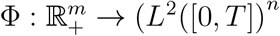, where ℝ_+_ denotes the nonnegative real numbers. We assume that Φ has a Fréchet derivative, i.e., the mapping 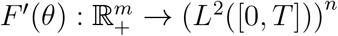, is injective, thus the forward problem (9) has a unique solution *x* for a given *θ*. The Fréchet derivative of Φ, denoted by Φ^′^, results to be the usual derivative for the system (8) since the domain and range of Φ^′^ are finite dimensional spaces. Usually, not all states of the system can actually be directed measured, i.e., the data consists of measurements of some state variables at a discrete set of points *t*_1_, …, *t*_*k*_, e.g. in epidemiology, these data consist of number of cases of confirmed infected people. This defines a linear observation mapping from state variables to data Ψ : (*L*^2^([0, *T*]))^*n*^ : → ℝ^*s×k*^, where *s* ≤ *n* is the number of observed variables and *k* is the number of sample points. Let *F* : ℝ^*m*^ → ℝ^*s×k*^ be defined by *F* (*θ*) = Ψ(Φ(*θ*)), called the forward problem. The inverse problem is formulated as a standard optimization problem

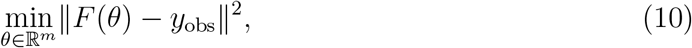

such that *x* = Φ(*θ*) holds, with *y*_obs_ is the data which has error measurements of size *η*. Problem (9) may be solved using numerical tools to deal with a nonlinear least-squares problem or the Landweber method or the combination of both. We implement Bayesian inference to solve the inverse problem (10) in this manuscript. From the Bayesian perspective, all state variables *x* and parameters *θ* are considered as random variables and the data *y*_obs_ is fixed. For random variables *x, θ*, the joint probability distribution density of data *x* and parameters *θ*, denoted by *π*(*θ, x*), is given by *π*(*θ, x*) = *π*(*x*|*θ*)*π*(*θ*), where *π*(*x*|*θ*) is the conditional probability distribution, also called the likelihood function, and *π*(*θ*) is the prior distribution which involves the prior information of parameters *θ*. Given *x* = *y*_obs_, the conditional probability distribution *π*(*θ*|*y*_obs_), called the posterior distribution of *θ* is given by the Bayes’ theorem:

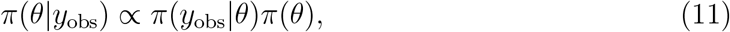

If additive noise is assumed:

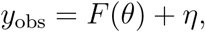

where *η* is the noise due to discretization, model error and measurement error. If the noise probability distribution *π*_*H*_ (*η*) is known, *θ* and *η* are independent, then

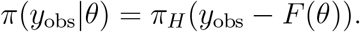

All the available information regarding the unknown parameter *θ* is codified into the a prior distribution *π*(*θ*), it specifies our belief in a parameter before observing the data. All the available information regarding the way of how was obtained the measured data is codified into the likelihood distribution *π*(*y*_obs_|*θ*). This likelihood can be seen as an objective or cost function, as it punishes deviations of the model from the data. To solve the associated inverse problem (11), one may use the maximum a posterior (MAP)

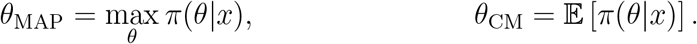

We used the data set 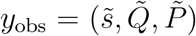, which correspond to the Suspects, diagnosed Sick cases and the Deceases, respectively. We mention that we have not used the data column corresponding to the Recovery people here because in a big range (from the beginning) of days this data was not been collected. A Poisson distribution with respect to the time is typically used to account for the discrete nature of these counts. However, the variance of each component of the data set *y*_obs_ is larger than its mean, which indicates that there is over-dispersion of the data. Thus, a more appropriate likelihood distribution is to use the Negative Binomial, since it has an additional parameter that allows the variance to exceed the mean [8, 17, 52]. In fact the Negative Binomial is a mixture of Poisson and Gamma distributions, where the rate parameter of the Poisson distribution itself follows a Gamma distribution [25, 52]. We mention there exists different mathematical expressions for the Negative Binomial depending on the author or source, they are equivalent. Because of this multiple representation of the NB in the literature, one must assure to use the NB distribution accordingly to the source. Here, we have used the following expression for the Negative Binomial distribution

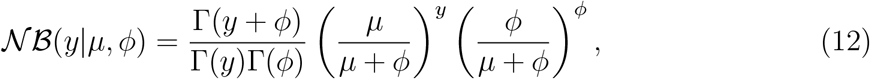

where *µ* is the mean of the random variable *y ∼ 𝒩 ℬ* (*y µ, ϕ*) and *ϕ* is the overdispersion parameter, i.e.,

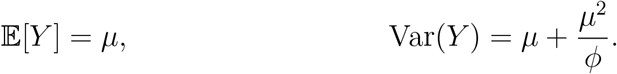

We recall that Poisson distribution has mean and variance equal to *µ*, so *µ*^2^*/ϕ >* 0 is the additional variance of the negative binomial with respect to the Poisson distribution. Therefore, the inverse of the parameter *ϕ* controls the overdispersion, this is important when selecting its support for parameter estimation. Also, there exists alternative forms of the Negative Binomial distribution. In fact, we have used the first option *neg*_*bin* of the Negative Binomial distribution of Stan [19]. We acknowledge that some scientists have had success with the second alternative representation of the NB distribution [31]. We assume independent Negative Binomial distributed noise *η*, i.e., all dependency in the data is codified into the contact tracing model. In other words, the positive definite noise covariance matrix *η* is assumed to be diagonal. Therefore, using Bayes formula, the likelihood is

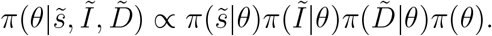

As mentioned above, we approximate the likelihood probability distribution corresponding to Suspects, Diagnosed cases, and Deaths with a Negative Binomial distribution

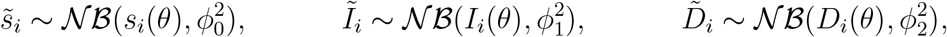

where the index *i* denotes the number time, in our case the number of days and *ϕ*_0_, *ϕ*_1_ and *ϕ*_2_ are the parameters corresponding to the overdispersion parameter of the Negative Binomial distribution (12), respectively of each data component. For independent observations, the likelihood distribution *π*(*y*|*θ*), is given by the product of the individual probability densities of the observations

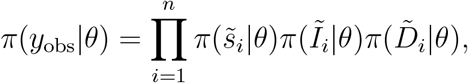

where the mean *µ* of the negative binomial distribution 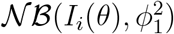, is given by the solution *I*(*t*) of the model (8) at time *t* = *t*_*i*_. Analogously, the mean for the negative binomial distributions 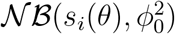 and 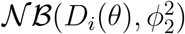 are the solutions *s*(*t*) and *D*(*t*) of (8) at time *t*_*i*_, respectively. For the prior distribution, we select LogNormal distribution for the *β* parameter and Uniform distributions for the rest of parameters to estimate: *q, δ, α, γ, σ, s*_0_, *E*_0_, *I*_0_, *Q*_0_. The hyperparameters and their support corresponding to all the distributions of the parameters to estimate are given on table.

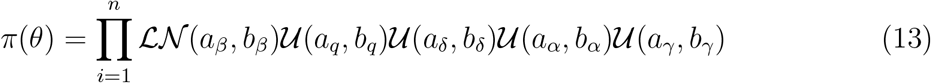

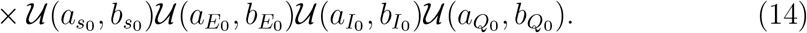

The posterior distribution *π*(*θ*|*y*_obs_) given by (11) does not have an analytical closed form since the likelihood function, which depends on the solution of the nonlinear SsEAIRD model, does not have an explicit solution. Then, we explore the posterior distribution using the Stan Statistics package [19], general purpose Markov Chain Monte Carlo Metropolis-Hasting (MCMC-MH) algorithm to sample it, the package *t-walk* [24]. Both algorithms generate samples form the posterior distribution *π*(*θ*|*y*_obs_) that can be used to estimate marginal posterior densities, mean, credible intervals, percentiles, variances, etc. We refer to [35] for a more complex MCMC MH algorithms. The dataset in [1] contains the information regarding the number of diagnosed cases, deaths, and suspects. Figures 6-8 show the results of forecasting the disease using the Stan package. Table 2 shows the parameter estimated using the Stan package with the quantiles 2.5%, 25%, 50%, 75%, 97.5%.

We perform 20000 iterations, with 10000 of them as a burnin. We have used the interface in Python (PyStan). We have used the Hamilton Monte Carlo and No-U-Turn Sampler (NUTS) algorithms, obtaining similar performance. We point out that using Automatic Differentiation Variational Inference (ADVI) is much faster than the previous algorithms mentioned, with also very similar results. Figures 9 and the right column of figure 7 show corresponding results using the *t-walk* package (the Python version of it). We performed 600000 iterations with 300000 of them as burnin. Using both packages, we have done predictions until the day 240, meaning October 16th. Some future work will correspond to analyze the identifiability of the parameters of model (8), as suggested in [22, 49, 57], specifically the *ρ* parameter since this parameter is multiplied by the period of incubation of the disease, *α*, thus, estimating both parameters simultaneously may lead to nonidentifiability difficulty. In this work, we have assumed the value for the period of incubation of the disease.

**Table 1:** Parameters of the contact tracing model (8).

| Parameter | Description | Value |
| --- | --- | --- |
| $\beta_s$ | transmission rate of the disease by simptomatics | to be estimated |
| $\beta_a$ | transmission rate of the disease by asimptomatics | to be estimated |
| $\rho$ | the fraction of asymptomatics/symptomatics | to be estimated |
| $1/\gamma$ | recovery period (days) | to be estimated |
| $\sigma$ | death rate by the disease | to be estimated |
| $q$ | fraction of Suspects | to be estimated |
| $1/\tau$ | period of quarantined (days) | 14 |
| $1/\alpha$ | latent period (days) | 5.1 |
| $E_0$ | initial condition for exposed class $E(0)$ | to be estimated |
| $A_0$ | initial condition for asymptomatic class $A(0)$ | to be estimated |
| $I_0$ | initial condition for symptomatic class $I(0)$ | to be estimated |

**Table 2:** Parameters of the contact tracing model (8).

|  | mean | 2.5% | 25% | 50% | 75% | 97.5% |
| --- | --- | --- | --- | --- | --- | --- |
| $\beta_s$ | 0.222620 | 0.044925 | 0.128393 | 0.211603 | 0.312559 | 0.433075 |
| $\beta_a$ | 0.329556 | 0.299295 | 0.323674 | 0.332539 | 0.338223 | 0.344950 |
| $\rho$ | 0.997225 | 0.996923 | 0.997114 | 0.997219 | 0.997329 | 0.997565 |
| $\gamma$ | 0.190190 | 0.167331 | 0.186027 | 0.192736 | 0.196920 | 0.199693 |
| $\sigma$ | 0.102499 | 0.089572 | 0.097630 | 0.102195 | 0.107137 | 0.116798 |
| $q$ | 0.056694 | 0.019540 | 0.027548 | 0.040514 | 0.066996 | 0.193501 |
| $E_0$ | 11780.966835 | 7526.224012 | 10093.482221 | 11627.748681 | 13273.949132 | 17010.362332 |
| $A_0$ | 7182.017997 | 4917.603144 | 6872.829362 | 7445.705247 | 7765.232788 | 7977.776073 |
| $I_0$ | 1.220856 | 0.130659 | 0.540569 | 0.970296 | 1.627982 | 3.748581 |
| $\phi_0$ | 4.048778 | 3.175233 | 3.712320 | 4.025514 | 4.363766 | 5.062059 |
| $\phi_1$ | 2.589235 | 2.060252 | 2.384828 | 2.576913 | 2.778927 | 3.192292 |
| $\phi_2$ | 2.389033 | 1.868520 | 2.191568 | 2.373326 | 2.573794 | 2.989858 |

**Figure 6:**
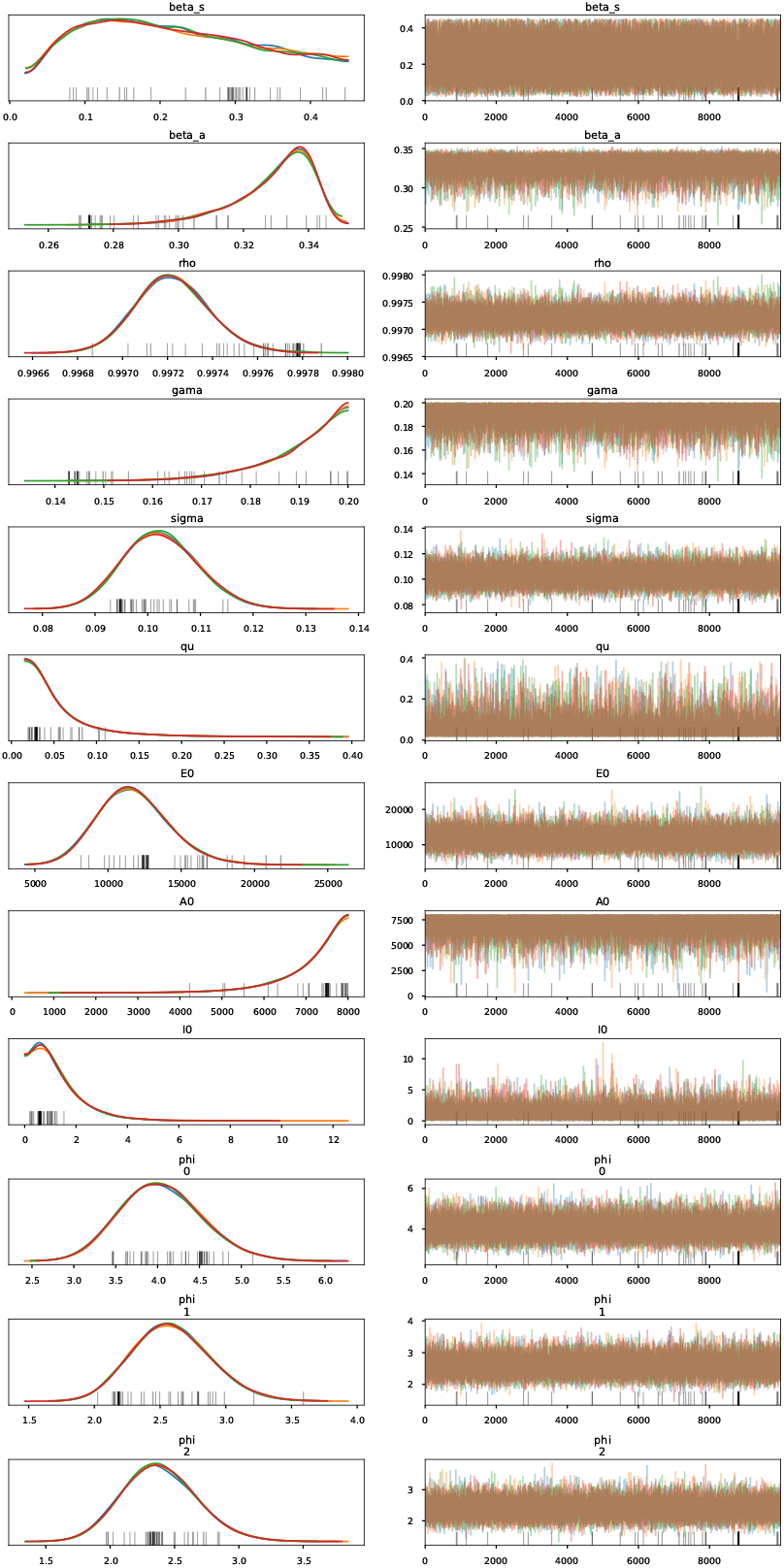
Credible intervals of parameters of model (8) within 95% Highest-Posterior Density (HPD)

**Figure 7:**
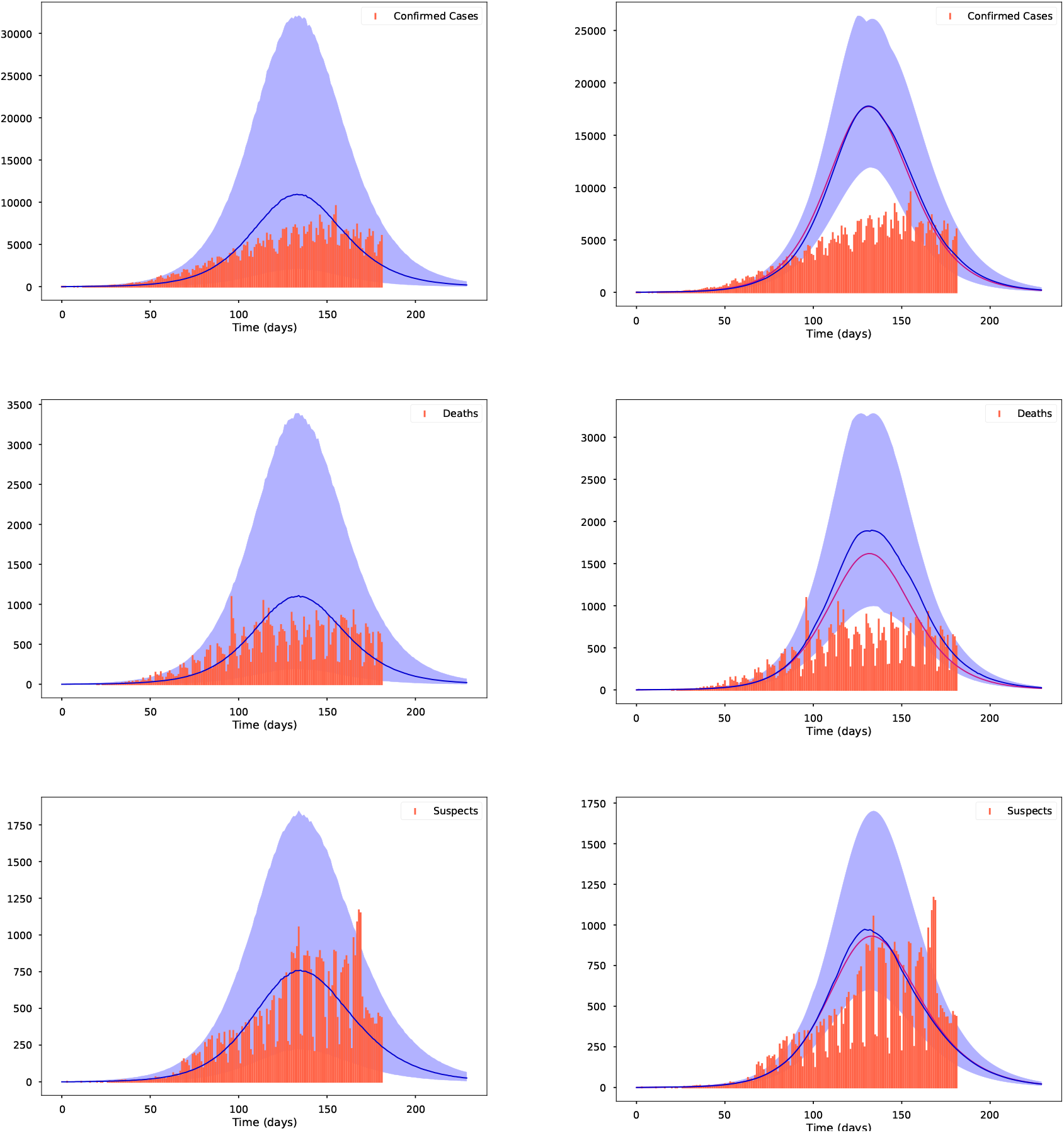
Left column from top to bottom: The fit for the diagnosed cases, for the deceases and for the suspects using the Stan package [19]. Right column from top to bottom: The fit for the diagnosed cases, for the deceases and for the suspects using the package *t-walk* package [24]. The red bars represent the observed data, the blue solid lines represent the medians and the shaded area represent the %95 probability bands for the expected value for the state variables: Infecteds, Deaths and Suspects, respectively. The solid purple line on the left column is the Maximum A Posteriori (MAP) curve.

**Figure 8:**
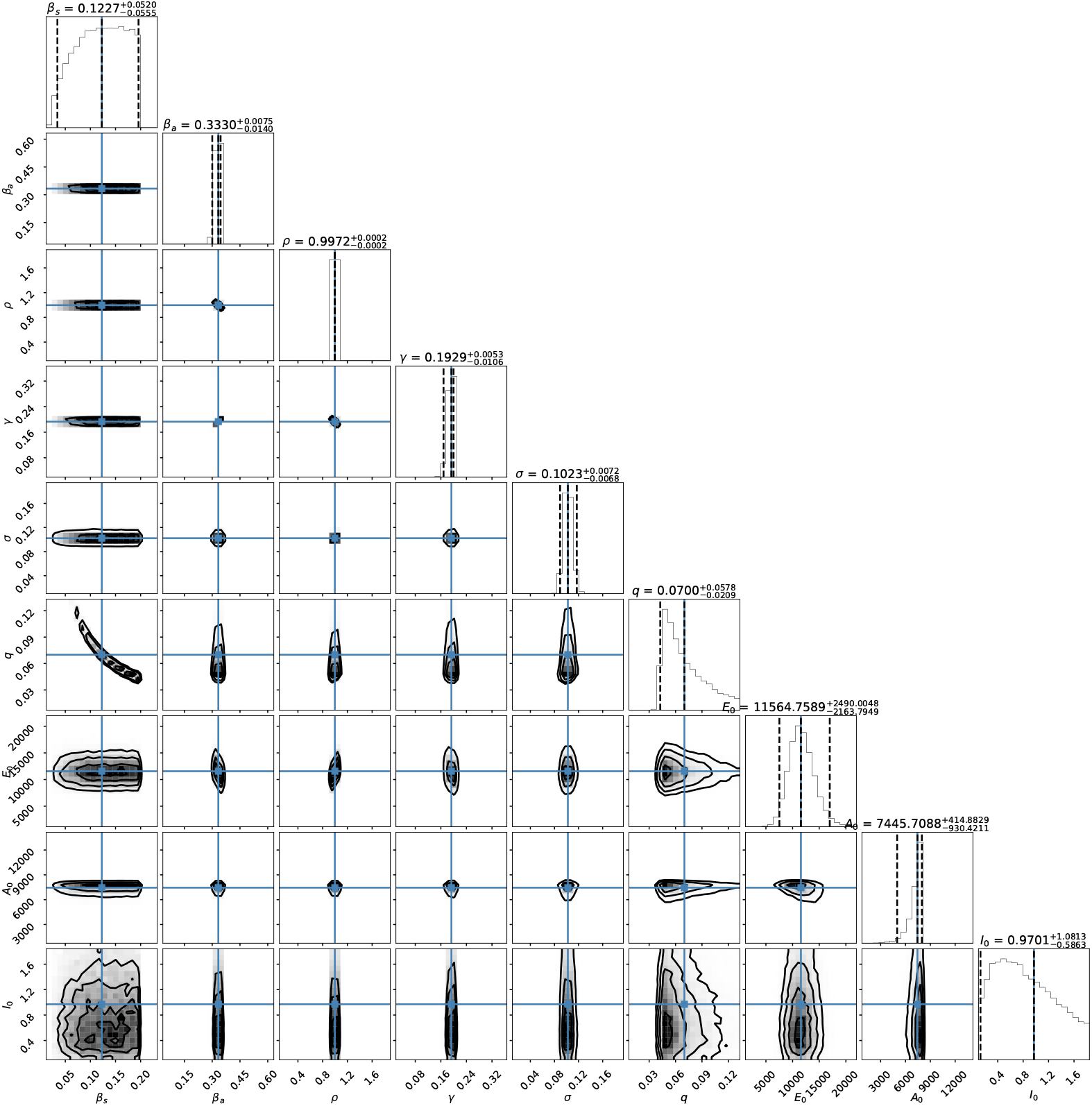
Joint probability density distributions of the estimated parameters within 95% (HPD). The blue lines represent the medians.

**Figure 9:**
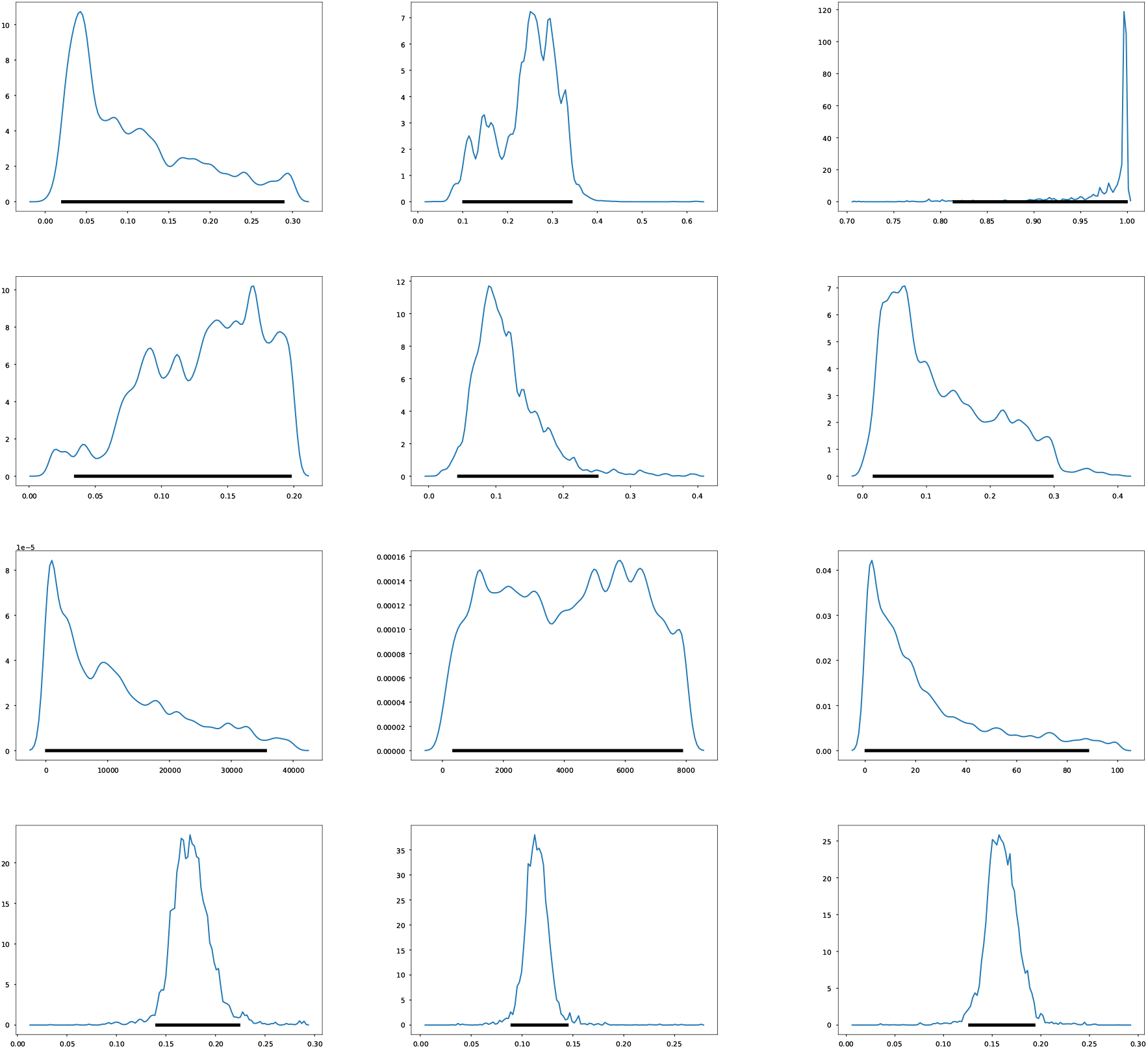
Credible intervals for the estimated parameters within %95 of HPD. Top row from left to right, the parameters: *β*_*s*_, *β*_*a*_, *ρ*. Middle row from left to right: *γ, σ, q*. Middle row from left to right: *E*_0_, *A*_0_, *I*_0_. Bottom row from left to right: *ϕ*_0_, *ϕ*_1_, *ϕ*_2_

## 4 Clinical analysis with Machine Learning

In this section, we describe the comorbidity associated with coronavirus in Mexico using the data set [1]. We have performed Machine Learning techniques on it as follows. Firstly, we implemented a predicted classifier for the kind of patient will be, a person already diagnosed with coronavirus and got one or more of the most relevant chronic diseases (hypertension or diabetes). We have used predictors methods in Machine Learning such as Logistic Regression, Decision Tree, and K-Neighbors classifiers, the naive Bayes (Bernoulli), even the powerful ones XGBoost and Random Forest through the SciKit-learn package. Figure 10 shows the covariance matrix of the most relevant chronic diseases with respect to the two types of patient: outpatient or hospitalized one. We can see in this figure that the most relevant chronic diseases with respect to the type of patient(outpatient or hospitalized) who has been diagnosed with coronavirus in Mexico are hypertension and diabetes. Table 3 shows the contingency table of these two chronic diseases with respect to to the type of patient. Figure 11 shows the relation in percent between outpatients and hospitalized patients. Figure 10 shows the confusion matrix result. We can add more characteristics like Age(range) to obtain more true negative cases since the differences in proportion of outpatient and hospitalized decreases. Next, instead of considering the type of patient (outpatient and hospitalized), we consider if the patient survives or deceases once diagnosed with coronavirus. Figure 10 shows the covariance matrix of the most relevant chronic diseases with respect to the two types of patient: survived or deceased. One can see in this figure that the most relevant chronic diseases with respect to the survival of a person who has been diagnosed with coronavirus in Mexico are hypertension and diabetes. Figure 14 shows the relation in percent between outpatients and hospitalized patients. Figure 15 shows the confusion matrix result. Adding more characteristics like age (range), one obtains similar results to figure 15, i.e., one obtains zero true negative predictions. We remind that false negative and false positive are the two type of errors of rejecting the hypothesis when it was actually true and accepting the hypothesis when it was actually false. Under different circumstances, one type of error may be more critical than the other. For example, diagnosis of cancer would rather accept false positives than false negatives. The main difficulty in trying to predict if a person will survive assuming that he has got either hypertension or diabetes is the rather unbalanced proportion between the two classes: survived and deceased. Unbalanced data is assumed with a category less than 20 percent. The lethality of coronavirus in the world is typically not greater than 15 percent.

**Table 3:**
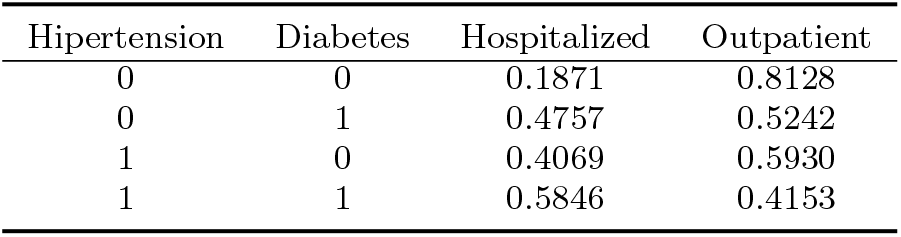
Patient type

**Table 4:** Survival patient

| Hipertension | Diabetes | Deceased | Survived |
| --- | --- | --- | --- |
| 0 | 0 | 0.0626 | 0.9374 |
| 0 | 1 | 0.2068 | 0.7931 |
| 1 | 0 | 0.1936 | 0.8063 |
| 1 | 1 | 0.3035 | 0.6964 |

**Figure 10:**
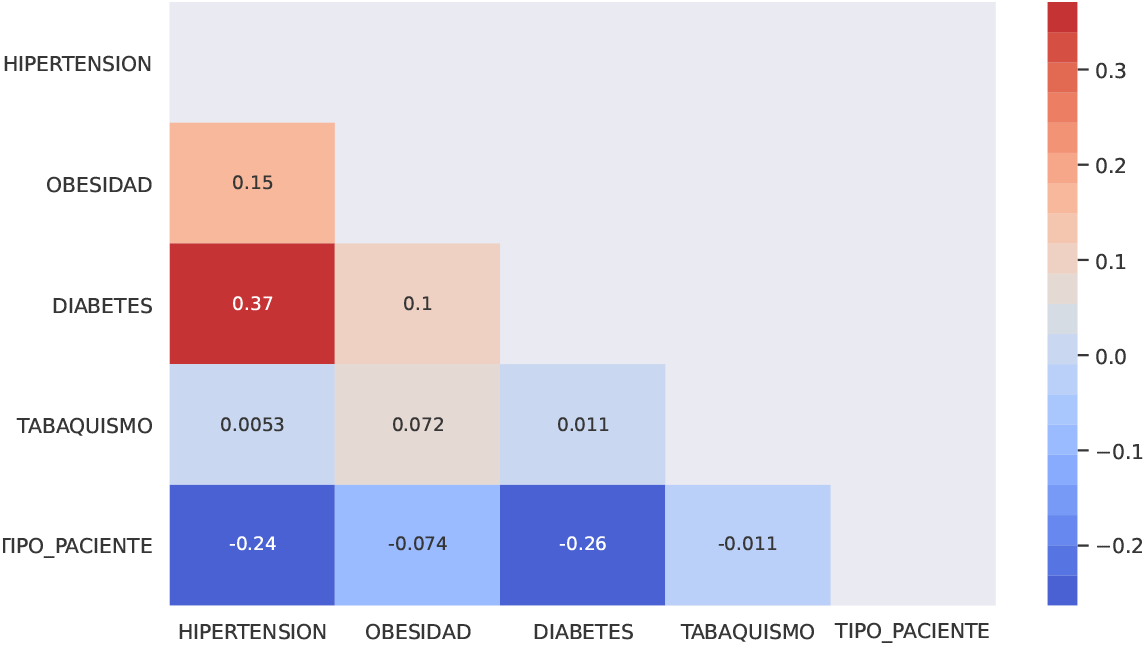
Covariance matrix of the most relevant chronic disaseses in Mexico with respect to the type of patient

**Figure 11:**
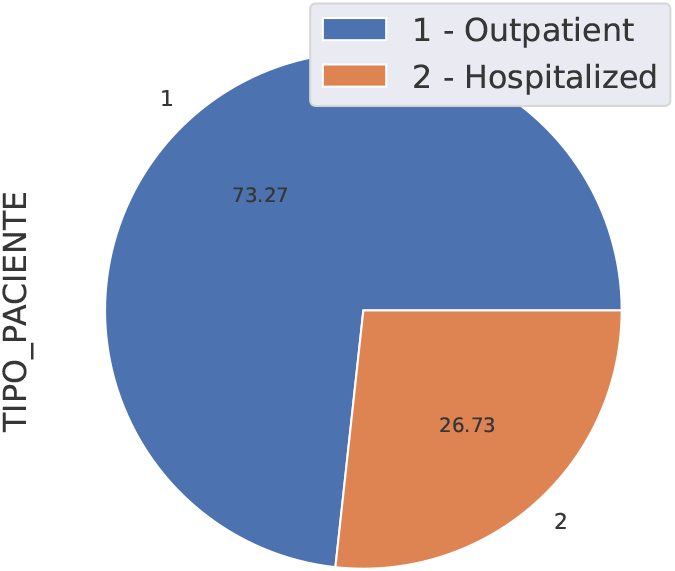
Relation outpatient-hospitalized

**Figure 12:**
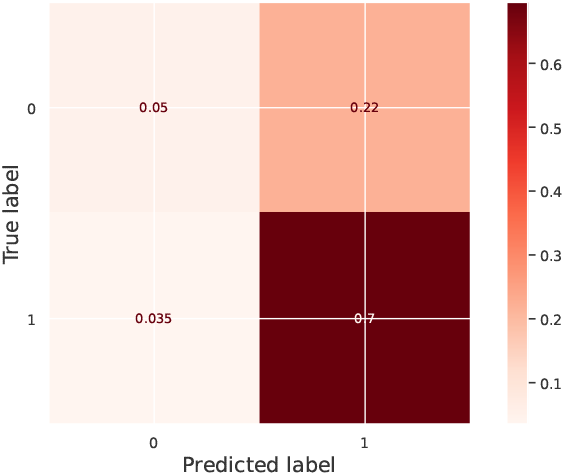
Confusion matrix

**Figure 13:**
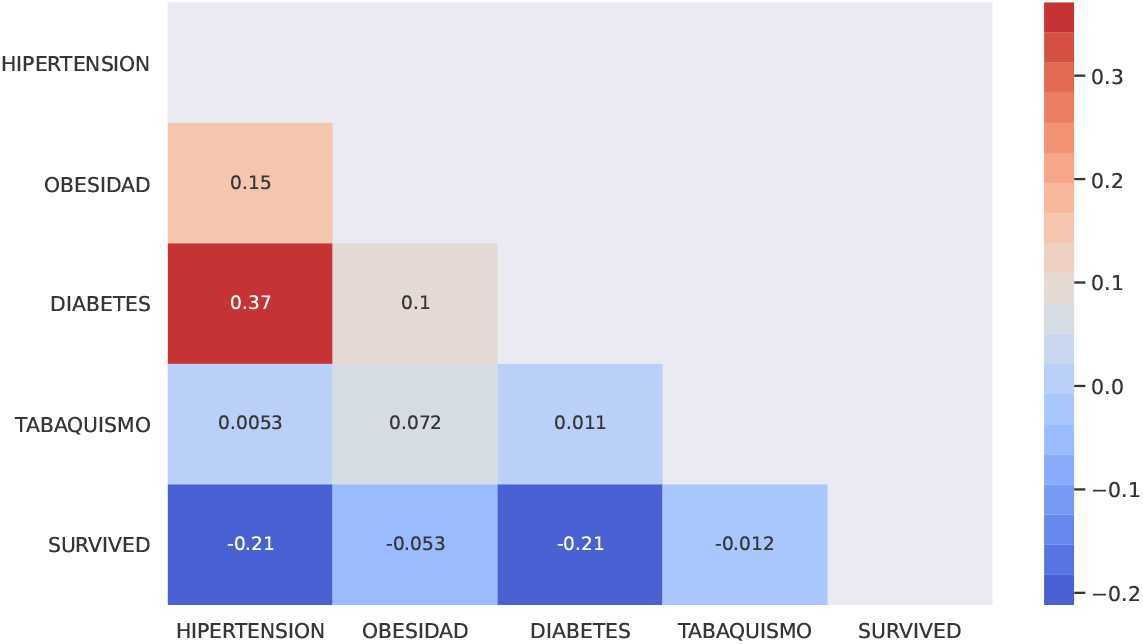
Covariance matrix of the most relevant chronic disaseses in Mexico with respect to the type of patient

**Figure 14:**
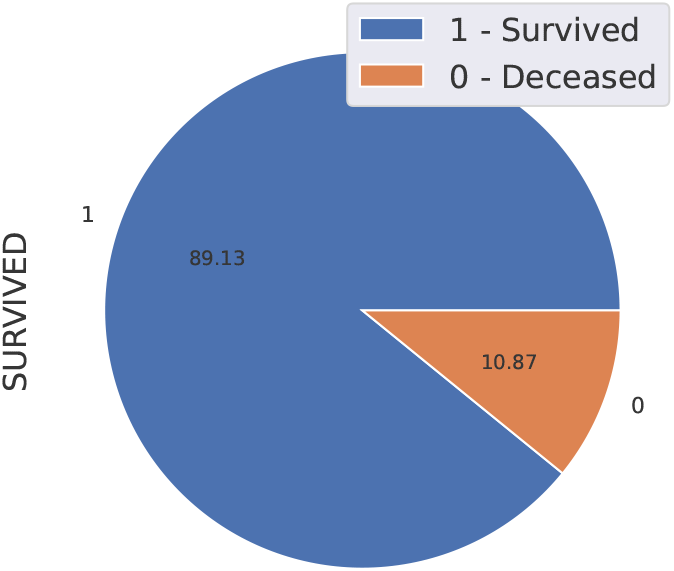
Relation survived-deceased

**Figure 15:**
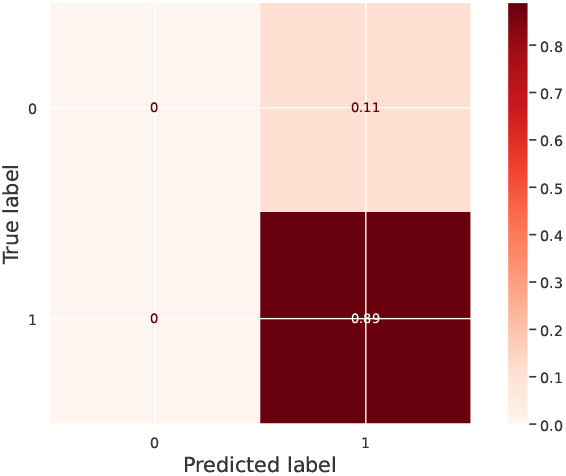
Confusion matrix

As it can be seen in 15, the true positives are very high but the prediction of true negatives are zero. We propose two options to deal with this difficulty. First, we have created a naive Bayes Multi-variate Bernoulli algorithm from scratch as suggested in [51]. This algorithm was originally proposed as an anti-spam email filter. Analogous to their description to classify spam emails, a person with vector *x* = ⟨*x*_1_, …, *x*_*m*_ ⟨,i.e., with multiple features but each one is assumed to be a binary-valued variable. In case of comorbidity, *x* represents the types of diseases. The decision rule for Bernoulli naive Bayes is based on the probability that a vector *x* belongs in category *c*:

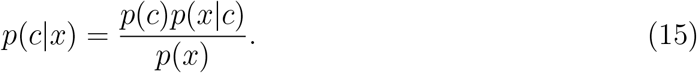

Since the denominator does not depend on the category, NB classifies each “message” in the category that maximizes the numerator in (15), i.e., *p*(*c*)*p*(*x*|*c*). In this case of “spam filter”, this is equivalent to classifying a message as spam whenever:

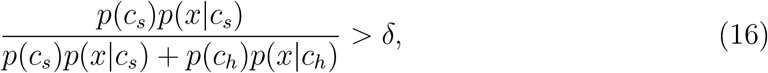

with *δ* = 0.5, where *c*_*h*_ and *c*_*s*_ denote the ham and spam categories. Here is the important part doing this algorithm from scratch, that we can vary *δ* to obtain more true negatives at the expense of true positives, or viceversa. In our case, we increased the true negatives, the number of true positives are very high using whatever classifier mentioned. Doing this, we can tune the threshold number of acceptance on the following formula 16. We selected *δ* = 0.45 (instead of 0.5) and obtained the following confusion matrix

As it can be seen in figure 16, the percent of true negatives has increased approximately to 2.6, also the false negative has decreased, although the false negative has increased too.

Secondly, we propose to use the Synthetic Minority Oversampling Technique (SMOTE) function to balance the minority class (people who passed away due to coronavirus). SMOTE briefly consists of synthesizing elements for the minority class, based on those that already exist. It works randomly picking a point from the minority class and computing the k-nearest neighbors for this point. The sysnthetic points are added between the chosen point and its neighbors. Figure 17 shows the result using the SMOTE technique. Another perspective in this comorbidity analysis may be to filter the people who were admitted to the hospital.

**Figure 16:**
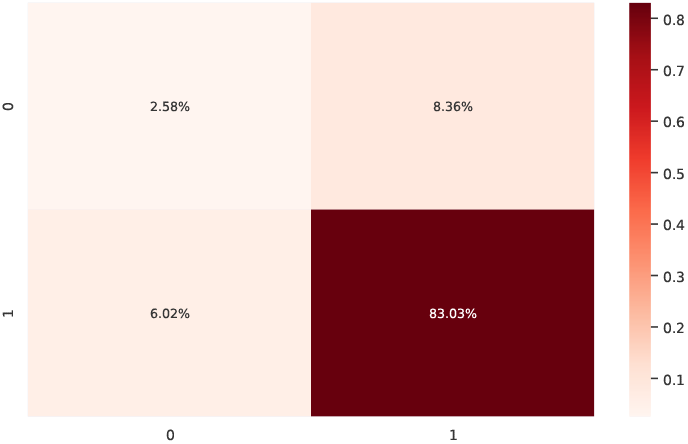
Confusion matrix applying Naive Bayes with threshold *δ* = 0.45

**Figure 17:**
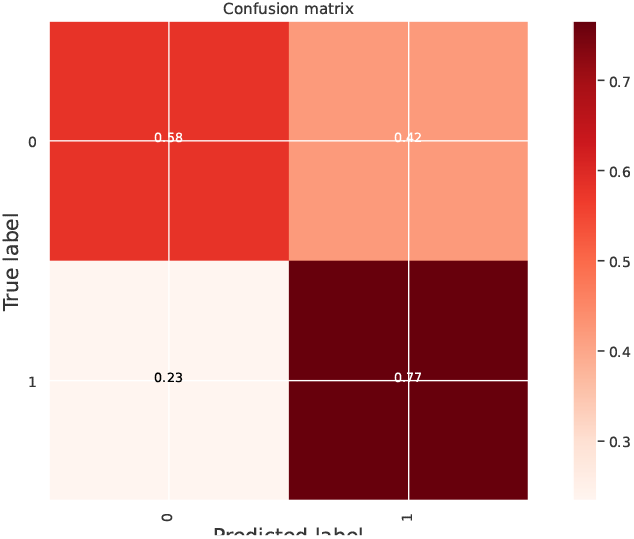
Confusion matrix using the SMOTE

## 5 Discussion and conclusions

In section 2, we present a formula to estimate the basic and effective reproductive numbers, *R*_0_ and *R*_*t*_, based on serological data using Bayesian inference and the Bayesian Learning Paradigm. We present the numerical results for the top five most affected regions in Mexico and for the whole country. We consider that these results may be reliable since the age range of scanned people for coronavirus, follows a Normal distribution around the mean age of the disease in Mexico, thus, the samples of our data are not unbiased.

In section 3, we present a forecast of transmission of the disease using Bayesian inference based on two softwares, the Stan package [19] and the *t-walk* package [24]. We show trace plots, credible intervals, bands projections with medians and a MAP curve (for the *t-walk* case) and the joint crosstab probability distributions given as a corner. From figure 7, we can see that the government of Mexico took some measures to control the transmission of the disease. Also, the value of the parameter, *ρ*, which refers to the proportion of symptomatics and asymptomatics was around .99, meaning that a big percent of asymptomatic for this disease.

In section 4, we explore some predictors of type of patient may be outpatient versus hospitalized and survived versus deceased using Machine Learning. Moreover, we present two methods to deal with unbalanced data as it is the case in coronavirus dataset in the world, specially for the case survived/deceased. Firstly, we propose to use the Naive Bayes method. Secondly, we propose to use the SMOTE technique. Both techniques have their pros and cons as explained in section 4.

## Data Availability

https://www.gob.mx/salud/documentos/datos-abiertos-152127

## Notes

### Competing Interest Statement

The authors have declared no competing interest.

### Funding Statement

No external funding was received

## References

[1] Covid-19 Mexico. https://coronavirus.gob.mx/datos/.1,2,2,3.2,3.2,4

[2] worldometers. https://www.worldometers.info/world-population/mexico-population/. 2

[3] Acunã Zegarra, M., Comas-García, A., Hernández-Vargas, E., Santana-Cibrian, M., and Velasco-Hernández, J. (2020). The SARS-CoV-2 epidemic outbreak: a review of plausible scenarios of containment and mitigation for Mexico. medRxiv.2, 3.1, 3.2

[4] Adhikari, R. and Bolitho, A. e. a. (2020). Inference, prediction and optimization of non-pharmaceutical interventions using compartment models: the PyRoss library. arXiv e-prints. 1

[5] Alavez-Ramirez, J. (2007). Estimacion de parámetros en ecuaciones diferenciales ordinarias: identificabilidad y aplicaciones a medicina. Revista electrónica de contenido matemático, 21. 3.2

[6] Alonso-Quesada, G., De la Sen, M., and Ibeas, A. (2017). On the discretization and control of an SEIR epidemic model with a periodic impulsive vaccination. Commun Nonlinear Sci Numer Simulat, 42:247–274. 1

[7] Anderson, R. and May, R. (1977). Population biology of infectious diseases: Part I. Nature, 280:361–367. 1

[8] Arguëdas, Y., Santana-Cibrian, M., and Velasco-Hernández, J. (2019). Transmission dynamics of acute respiratory diseases in a population structured by age. Mathematical Biosciences and Engineering, 16(6):7477–7493. 3.2, 3.2

[9] Bettencourt, L. and Ribeiro, R. (2008). Real Time Bayesian Estimation of the Epidemic Potential of Emerging Infectious Diseases. PlosOne, 3(5):e2185. 3.2

[10] Bliznashki, S. (2020). A Bayesian Logistic Growth Model for the Spread of COVID-19 in New York. medRxiv, 14(12). 1, 3.2

[11] Boersch-Supan, P., Ryan, S., and Johnson, L. (2017). deBInfer:Bayesian inference for dynamical models of biological systems in R. Methods in Ecology and Evolution, 8:511–518. 3.2

[12] Bolzoni, L., Bonacini, E., Soresina, C., and Groppi, M. (2017). Time-optimal control strategies in SIR epidemic models. Mathematical Biosciences, 292:86–96. 1

[13] Brauer, F., van den Driessche, P., and Wu, J. (2008). Mathematical epidemiology. Springer. 3.1

[14] Brown, G., Porter, A., Oleson, J., and Hinman, J. (2018). Approximate Bayesian computation for spatial SEIR(S) epidemic models. Spatial and Spatio’temporal Epidemiology, 24(10):2685–2697. 1, 3.2

[15] Browne, C., Gulbudak, H., and Webb, G. (2015). Modeling contact tracing in outbreaks with application to Ebola. Journal of Theoretical Biology, 384:33–49. 3.1

[16] Cao, C., Chen, W., Zheng, S., Zhao, J., Wang, J., and Cao, W. (2016). Analysis of Spatiotemporal Characteristics of Pandemic SARS Spread in Mainland China. BioMed Research International, 2016:889–917. 1

[17] Capistrán, M., Capella, A., and Christen, A. (2020). Forecasting hospital demand during COVID-19 pandemic outbreaks. arXiv e-prints. 3.1, 3.2, 3.2

[18] Capistrán, M., Moreles, M., and Lara, B. (2009). Parameter Estimation of Some Epidemic Models. The Case of Recurrent Epidemics Caused by Respiratory Syncytial Virus. Bulletin of Mathematical Biology, 71(8):1890–1901. 3.2

[19] Carpenter, B., Gelman, A., Hoffman, D., Goodrich, B., Betancourt, M., Brubaker, M., Guo, J., Li, P., and Riddell, A. (2017). Stan: A probabilistic programming language. Journal of Statistical Software, 76(1):1–32. 3.2, 3.2, 7, 5

[20] Chandra, V. (2020). Stochastic compartmental modelling of sars-cov-2 with approximate bayesian computation. medRxiv. 1

[21] Chatzilena, A., Leeuwen, E., Ratmann, O., Baguelin, M., and Demiris, N. (2019). Contemporary statistical inference for infectious disease models using Stan. Epidemics, 29. 1, 3.2

[22] Chowell, G. (2017). Fitting dynamic models to epidemic outbreak with quantified uncertainty: A primer for parameter uncertainty, identifiability, and forecasts. Infectious Disease Modelling, 2:379–398. 3.2, 3.2

[23] Chowell, G., Fenimore, P., Castillo-Garsow, M., and Castillo-Chavez, C. (2003). SARS outbreaks in Ontario, Hong Kong and Singapore: the role of diagnosis and isolation as a control mechanism. Journal of Theoretical Biology, 224:1–8. 1, 3.1

[24] Christen, J. and Fox, C. (2010). A general purpose sampling algorithm for continuous distributions (the t-walk). Bayesian Anal., 5:263–282. 2, 3.2, 7, 5

[25] Coly, S., Yao, A., Abrial, D., and Garrido, M. (2016). Disributions to model overdis-persed count data. Journal de la Societe Francaise de Statistique, 157(2):39–63. 3.2

[26] Dehning, J., Zierenberg, J., Spitzner, P., Wibral, M., Neto, J., Wilczek, M., and Priesemann, V. (2020). Inferring change points in the COVID-19 spreading reveals the effectiveness of interventions. Science, 369(10). 1

[27] Ferretti, L., Wymant, C., Kendall, M., Zhao, L., Nurtay, A., Abeler-Dörner, L., Parker, M., Bonsall, D., and Fraser, C. (2020). Quantifying sars-cov-2 transmission suggests epidemic control with digital contact tracing. Science, 368(6491). 3.1

[28] Flaxman, S., Mishra, S., and Gandy, A. e. a. (2020). Estimating the effects of non-pharmaceutical interventions on COVID-19 in Europe. Nature, 584:257–261. 1

[29] Gill, J. (2014). Bayesian Methods: A social and Behavioral Sciences Approach. Chapman & Hall/CRC, third edition edition. 2

[30] Greenhalgh, D. (1997). Hopf Bifurcation in Epidemic Models with a Latent Period and Nonpermanent Immunity. Mathl. Comput. Modelling, 25(2):85–107. 1

[31] Grinsztajn, L., Semenova, E., Margossian, C., and Riou, J. (2020). Bayesian workflow for disease transmission modeling in Stan. arXiv e-prints. 3.2, 3.2

[32] Heesterbeek, H., Anderson, R., Andreasen, V., Bansal, S., and Daniela, A. (2020). Modeling infectious disease dynamics in the complex landscape of global health. Science, 347(6227). 1

[33] Hethcote, H. (2000). The Mathematics of Infectious Diseases. SIAM REVIEW, 42(4):599–653. 3.1

[34] Hethcote, H., Zhien, M., and Shengbing, L. (2002). Effects of quarantine in six endemic models for infectious diseases. Mathematical Biosciences, 180:141–160. 3.1

[35] House, T., Ford, A., Lan, S., Bilson, S., Buckingham-Jeffery, E., and Girolami, M. (2016). Bayesian uncertainty quantification for transmissibility of influenza, norovirus and Ebola using information geometry. J.R.Soc. Interface, 13. 3.2

[36] Keeling, M. and Rohani, P. (2008). Modeling Infectious Diseases in humans and animals. Princeton University Press. 1, 2, 3.1, 3.1

[37] Kim, S., Kwon, H.-D., and Lee, J. (2016). Constrained optimal control applied to vaccination for influenza. Computers and Mathematics with Applications, 71:2313–2329. 1

[38] Kim, S., Lee, J., and Jung, E. (2017). Mathematical model of transmission dynamics and optimal control strategies for 2009 A/H1N1 influenza in the Republic of Korea. Journal of Theoretical Biology, 412:74–85. 1

[39] Kornienko, I., Paiva, L., and de Pinho, M. (2017). Introducing State Constraints in Optimal Control for Health Problems. Procedia Technology, 17:415–422. 1

[40] Korobeinikov, A. (2004). Lyapunov functions and global properties for SEIR and SEIS epidemic models. Mathematical Medicine and Biology, 21:75–83. 1

[41] Kwok, K., Tang, A., Wei, V., Park, W., Yeoh, E., and Riley, S. (2019). Epidemic Models of Contact Tracing: Systematic Review of Transmission Studies of Severe Acute Respiratory Syndrome and Middle East Respiratory Syndrome. Computational and Structural Biotechnology Journal, 17:186–194. 3.1

[42] Li, M. and Muldowney, J. (1995). Global stability for the SEIR model in epidemiology. Mathematical Biosciences, 125(4):155–164. 1

[43] Li, M. and Wang, L. (2002). Global stability in some SEIR epidemic models. In Mathematical Approaches for Emerging and Reemerging Infectious Diseases: Models, Methods, and Theory, number 2, pages 295–311. Springer New York. 1

[44] Lin, Q. and Zhao, S. e. a. (2020). A conceptual model for the coronavirus disease 2019 (COVID-19) outbreak in Wuhan, China with individual reaction and governmental action. International Journal of Infectious Diseases, 93:211–216. 1

[45] Lipsitch, M. and Cohen, T. e. a. (2020). Transmission dynamics and control of severe acute respiratory syndrome. Science, 300(5627). 1

[46] Liu, W., Hethcote, H., and Levin, S. (1987). Dynamical behaviour of epidemiological models with non-linar incidence rates. J. Math. Biol., 25:359–380. 1

[47] Liu, W., Levin, S., and Iwasa, Y. (1986). Influence of non-linear incidence rates upon the behaviour of SIRS epidemiological models. J. Math. Biol., 23:187–204. 1

[48] Luzyanina, T. and Bocharov, G. (2018). Markov chain Monte Carlo parameter estimation of the ODE compartmental cell growth model. Mathematical Biology and Bioinformatics, 13:376–391. 3.2

[49] Magal, P. and Webb, G. (2018). The Parameter Identification Problem for SIR Epidemic Models: Identifying Unreported Cases. Journal of Mathematical Biology, 77:1629–1648. 3.2

[50] McBryde, E., Gibson, G., Pettitt, A., Zhang, B., and McElwain, D. (2006). Bayesian Modelling of an Epidemic of Severe Acute Respiratory Syndrome. Bulletin of Mathematical Biology, 68:889–917. 1, 3.2

[51] Metsis, V., Androutsopoulos, I., and Paliouras, G. (2006). Spam filtering with Naive Bayes-Which Naive Bayes? 3rd Conf. on Email and Anti-Spam (CEAS), 347. 4

[52] Nayens, T. and Faes, C. and Molenberghs, G. (2012). A generalized Poisson-gamma model for spatially oversdispersed data. Spatial and Spatio-temporal Epidemiology, 3:185–194. 3.2

[53] Obadia, T., Haneef, R., and Pierre-Yves, B. (2012). The R0 package: a toolbox to estimate reproduction numbers for epidemic outbreaks. Medical Informatics and Decision Making, 12(147):1–9. 2

[54] Prieto, K. and Dorn, O. (2016). Sparsity and level set regularization for diffuse optical tomography using a transport model in 2D. Inverse Problems, 33(1):014001. 3.2

[55] Prieto, K. and Ibarguen-Mondragon, E. (2019). Parameter estimation, sensitivity and control strategies analysis in the spread of influenza in Mexico. Journal of Physics: Conference Series, 1408(1):012020. 3.2

[56] Roda, W., Varughese, M., Han, D., and Li, M. (2020). Why is it difficult to accurately predict the COVID-19 epidemic? Infectious Disease Modelling, 5:271–281. 1, 3.1

[57] Roosa, K. and Chowell, G. (2019). Assesing parameter identifiability in compartmental dynamic models using a computational approach: application to infectious disease transmission models. Theoretical Biology and Medical Modelling, 16(1). 3.2

[58] Rudas, T. (2008). Handbook of Probability: Theory and Applications. SAGE Publications. 2

[59] Shim, E., Tariq, J., Choi, W., Lee, Y., and Chowell, G. (2020). Transmission potential and severity of COVID-19 in South Korea. International Journal of Infectious Diseases, 93:339–344. 2

[60] Smirnova, A., DeCamp, L., and Liu, H. (2016). Inverse Problems and Ebola Virus Disease Using an Age of Infection Model, pages 103–121. Springer,Cham. 3.2

[61] Song, P., Wang, L., and Zhou, S. e. a. (2020). An epidemiological forecast model and software assessing interventions on COVID-19 epidemic in China. medRxiv. 1

[62] Stojanović, O., Leugering, J., Pipa, G., Ghozzi, S., and Ullrich, A. (2019). A Bayesian Monte Carlo approach for predicting the spread of infectious diseases. PLoS ONE, 14(12). 3.2

[63] Waltman, P. (1974). Deterministic threshold models in the theory of epidemics, volume 1. Springer-Verlag, New York. 1

[64] Zhou, C. (2020). Evaluating new evidence in the early dynamics of the novel coronavirus covid-19 outbreak in wuhan, china with real time domestic traffic and potential asymptomatic transmissions. medRxiv. 1

